# Associations of current and childhood socioeconomic status and health outcomes amongst patients with knee or hip osteoarthritis in a Mexico City family-practice setting

**DOI:** 10.1101/2022.04.02.22272992

**Authors:** Julio Pisanty-Alatorre, Omar Yaxmehen Bello-Chavolla, Eduardo Vilchis-Chaparro, María Victoria Goycochea-Robles

**Author notes:** Address for correspondence: Insurgentes sur 1194-503, Col Del Valle CP 03100 Mexico City.

## Abstract

**Objectives:** To examine the association of current and childhood socioeconomic status (SES) with patient-reported functional status, quality of life and disability in patients with knee osteoarthritis (OA)

**Methods:** We conducted a cross-sectional study amongst individuals seeking care for any medical reason in a primary care family-practice clinic in Mexico City. We included individuals with self-reported doctor-diagnosed arthritis and administered a survey using validated Spanish language versions of the Western Ontario and McMaster Universities Osteoarthritis Index (WOMAC), the Osteoarthritis of Lower Limbs and Quality of Life (AMICAL), and the Stanford Health Assessment Questionnaire-Disability Index (HAQ-DI). To estimate current and childhood SES, we and used a validated tool to estimate income quintile, as well as education level and occupation type, for both the patient and their parents.

**Results:** We recruited 154 patients and excluded 8 patients. Estimated income and education levels were correlated with WOMAC, AMICAL and HAQ-DI scores, and significant differences were found in all scores by occupation type. The association for estimated income and all scores remained significant independently of age, sex, BMI, and presence of diabetes or hypertension. Maternal education was best correlated with AMICAL scores, though its effect seemed largely mediated by its association with current SES measures.

**Conclusions:** Current and – to a lesser extent – childhood Socioeconomic Status impacts functional status, quality of life and disability amongst OA patients in Mexico City. Awareness of life-course SES can help identify patients at risk for worse outcomes.

## Introduction

Socioeconomic Status (SES) has attracted increasing interest as a determinant of both prevalence and outcomes for chronic health conditions, including osteoarthritis (OA) and other musculoskeletal disorders.(1,2) A large body of research shows the deleterious impact on health of low SES measured through the proxies of occupation, education and income, (3) which have been shown to be equal or better predictors of health outcomes than other compound measures of SES.(4)

Life-course epidemiology has identified lower SES during childhood as an important predictor of adult health, even independently of adult SES. In particular, the so-called pathway model proposes that low childhood SES sets individuals on a life trajectory in which a series of deleterious exposures are likely to occur, with SES at later stages of life mediating the harmful effect of low childhood SES.(5)

Various studies have examined the relationship between SES and OA prevalence, with the vast majority indicating higher prevalence rates amongst lower-SES groups.(6–8) Similarly, a growing number of studies are now painting a clear picture of a relationship between lower SES and worse OA outcomes.(9–12)

Similarly, a growing body of evidence shows an association between low childhood SES and increased prevalence of arthritis and of musculoskeletal pain (of which knee and hip OA probably play a large part). This relationship is partially mediated by adult SES.(13,14) Only one study has directly examined the relationship of childhood SES with arthritis outcomes. It found that low childhood SES is associated with higher disability and worse physical health amongst patients with arthritis (of whom most suffered from OA), after adjusting for current SES, age, sex, race, Body Mass Index (BMI) and comorbidities.(11) Few studies have examined the relationship of life course SES and OA outcomes in developing countries and, in particular, in Latin America, where exposure to low SES throughout the life-course is thought to be higher.(15)

The aim of this study was to examine the association of current and childhood SES with patient-reported functional status, quality of life and disability in a sample of patients with self-reported doctor-diagnosed primary knee or hip osteoarthritis seeking care in a family practice setting in Mexico City.

## 2. Methods

### 2.1 Study design and population

We conducted a cross-sectional study amongst individuals seeking care for any medical reason in a primary care family-practice clinic belonging to the Mexican Social Security Institute (IMSS) in Mexico City. We included adult patients with self-reported doctor-diagnosed knee or hip OA who signed informed consent. We excluded patients suffering from rheumatoid arthritis or other rheumatic conditions susceptible to causing secondary OA, as well as those who did not complete the general patient data questionnaire or withdrew consent after completion of questionnaires.

### 2.2 Measurements

We assessed functional status, quality of life and disability using validated Spanish language versions of the Western Ontario and McMaster Universities Osteoarthritis Index (WOMAC),(16) the Osteoarthritis of Lower Limbs and Quality of Life (AMICAL),(17) and the Stanford Health Assessment Questionnaire-Disability Index (HAQ-DI).(18)

We asked patients to complete a general data questionnaire including questions on level of education and occupation type for both themselves and their parents. To measure income, we administered Gutiérrez *et al*’s questionnaire, which has been shown to reliably estimate income quintile in the general Mexican population. (19)

### 2.3 Statistical Analysis

We used descriptive statistics for all variables. For ordinal variables education level, paternal and maternal education, and income quintile, we measured the correlation with the patient-reported outcomes by calculating Spearman coefficients (“rho”). For categorical variables occupation type, paternal and maternal occupation we used Kruskal-Wallis tests to identify differences in health outcomes, with post-hoc pairwise comparisons using Dunn tests with Bonferroni correction for comparisons against a specified group (Managerial office work).

In a post-hoc exploratory analysis, we dichotomized maternal education (high=elementary school or higher vs. low), and income (high= 4^th^-5^th^ income quintiles vs low). We then used these dichotomized variables to construct multiple linear regression models, entering the variables first separately and then concurrently, adjusting for age, sex, BMI and presence of Diabetes and Hypertension. Finally, we constructed SES trajectories defined by maternal education and current income, and constructed multiple linear regression models with this compound variable. All statistical analyses were done using R software version 4.1.0, with a chosen significance threshold value of p<0.05.

## Results

### Participants

We recruited 154 patients and excluded 8 patients (5 with rheumatoid arthritis, 1 with undifferentiated arthritis and 2 who withdrew consent), leaving a total of 146 patients for final analysis. Relevant patient characteristics are shown in Table 1. A majority (80%) of patients were female, with a mean age of 69.4 years. Because only 3% of patients had an estimated income in the lowest quintile, we grouped them with patients with estimated income in the second-lowest quintile.

**Table 1.** Patient characteristics.

|  | **Mean ± SD** |  |
| --- | --- | --- |
| **Age, years** | 69.5 ± 10.2 |  |
| **Body Mass Index, kg/m<sup>2</sup>** | 28.4 ± 4.9 |  |
| **WOMAC Score** | 44.6 ± 20.3 |  |
| **AMICAL Score** | 235.6 ± 73.8 |  |
| **HAQ-DI Score** | 1.14 ± 0.65 |  |
|  | **n** | **%** |
| **Sex, female** | 117 | 80 |
| **Smoking history, pack years** |  |  |
| 0 | 93 | 64 |
| >0, ≤ 5 | 25 | 17 |
| >5, ≤ 10 | 7 | 5 |
| >10, ≤ 15 | 4 | 3 |
| ≥15 | 17 | 11 |
| **Diabetes** | 41 | 28 |
| **Hypertension** | 76 | 52 |
| **Education** |  |  |
| Less than elementary (<6 years) | 25 | 17 |
| Elementary school (6 years) | 29 | 20 |
| Middle-school (9 years) | 31 | 21 |
| High-school (9 years) | 39 | 27 |
| College level or above | 22 | 15 |
| **Occupation type** |  |  |
| Manual labor | 67 | 47 |
| Non-managerial office work | 30 | 21 |
| Managerial | 22 | 15 |
| Home | 25 | 17 |
| **Estimated income quintile** |  |  |
| I or II | 21 | 14 |
| III | 30 | 21 |
| IV | 31 | 21 |
| V | 64 | 44 |
| **Paternal education** |  |  |
| Less than elementary (<6 years) | 48 | 36 |
| Elementary school (6 years) | 55 | 41 |
| Middle-school (9 years) | 12 | 9 |
| High-school (9 years) | 7 | 5 |
| College level or above | 13 | 9 |
| **Maternal education** |  |  |
| Less than elementary (<6 years) | 66 | 45 |
| Elementary school (6 years) | 53 | 36 |
| Middle-school (9 years) | 7 | 5 |
| High-school (9 years) | 11 | 8 |
| College level or above | 4 | 3 |
| **Paternal occupation** |  |  |
| Manual labor | 111 | 79 |
| Non-managerial office work | 10 | 7 |
| Managerial | 20 | 14 |
| **Maternal occupation** |  |  |
| Manual labor | 36 | 25 |
| Non-managerial office work | 9 | 6 |
| Managerial | 4 | 3 |
| Home | 94 | 66 |

SD: Standard Deviation; kg/m<sup>2</sup>: kilograms per meter squared; WOMAC: Western Ontario and MacMaster Universities Osteoarthritis Index; AMICAL: Osteoarthritis of Lower Limbs and Quality of Life; Health Assessment Questionnaire Disability Index

### Current and childhood SES and outcomes

Analysis of current SES variables showed that all three outcome scores were inversely correlated with both estimated current income quintile and higher education level. In general, there were significant differences in all three outcome scores according to patients’ occupation. However, subgroup analysis showed that compared to patients with managerial office work, only patients dedicated to manual labor had significantly higher scores (Figure 1).

**Figure 1.**
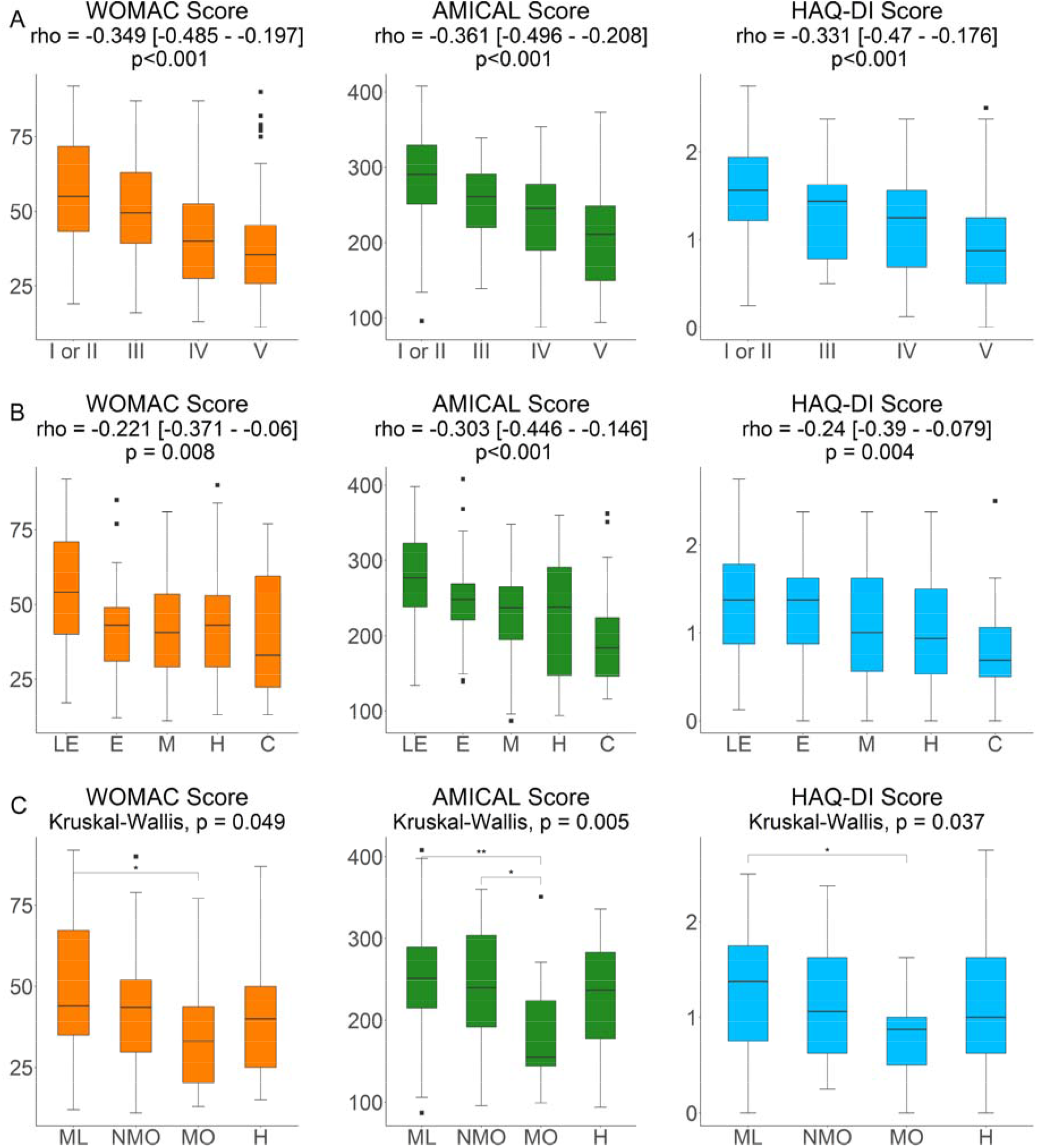
WOMAC, AMICAL and HAQ-DI Scores by current SES variables. Outcome scores by *A)* Estimated Income Quintile *B)* Education level and *C)* Occupation type. Spearman analysis for trend was conducted for *A* and *B*. Kruskall-Wallis testing was conducted for *C*, with post-hoc Dunn tests with Bonferroni correction. WOMAC: Western Ontario and MacMaster Universities Osteoarthritis Index; AMICAL: Osteoarthritis of Knee and Hip and Quality of Life; HAQ-DI: Stanford Health Assessment Questionnaire Disability Index. For education level, LE: less than elementary (<6 yr), E: elementary (6 years), M: Middle-school (9 years), H: High-school (12 years), C: College-level or higher (>12 years). For occupation type, H: Home, ML: Manual Labor, NMO: Non-managerial office work, MO: Managerial office work. *Significant at the p<0.05 level; ** Significant at the p<0.01 level

Regarding childhood SES, both maternal and paternal education were inversely correlated with AMICAL scores, but not WOMAC or HAQ-DI scores. There were significant differences in AMICAL, but not WOMAC or HAQ-DI scores, by maternal occupation type, and no significant differences in any score by paternal occupation type (Supplementary Figure 1)

### Interaction between current and childhood SES and with co-variates

Linear regression models demonstrated that current income significantly affects WOMAC, AMICAL and HAQ-DI scores independently of age, sex, BMI, and presence of diabetes or hypertension (Figure 2A). After adjusting for these same variables, maternal education had no effect on any outcome scores (Supplmentary Figure 2A).

**Figure 2.**
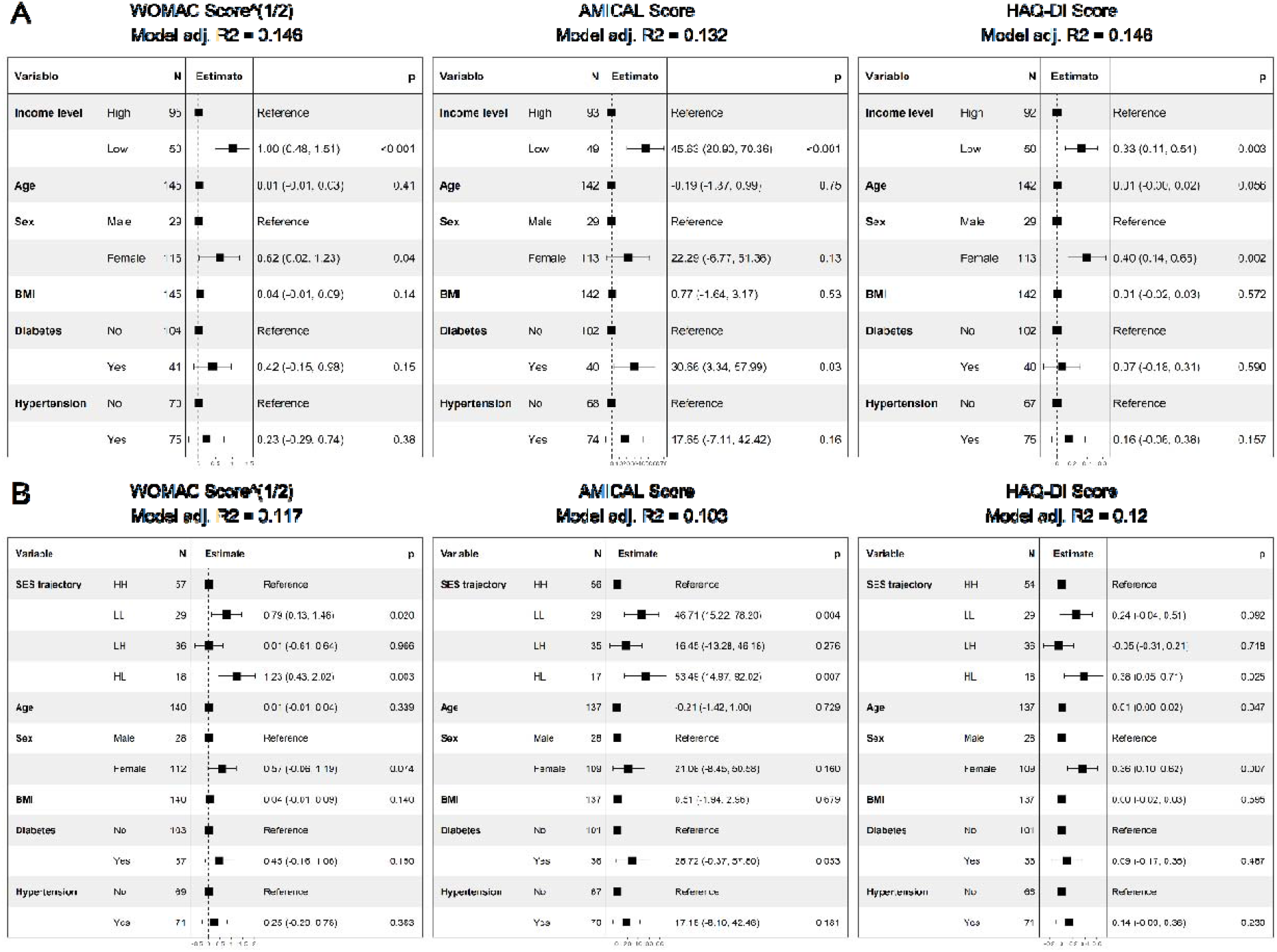
Linear regression models of effect of current income level and SES Trajectory on WOMAC, AMICAL and HAQ-DI scores. A) Beta coefficients of regression models only including current income level. B) Beta coefficients of regression models including Socio-Economic Status (SES) Trajectory HH: High Maternal Education/ High Income; LL: Low Maternal Education/ Low Income; LH: Low Maternal Education/ High Income; HL: High Maternal Education/ Low Income WOMAC: Western Ontario and MacMaster Universities Osteoarthritis Index; AMICAL: Osteoarthritis of Knee and Hip and Quality of Life; HAQ-DI: Stanford Health Assessment Questionnaire Disability Index; SES: Socio-Economic Status; BMI: Body Mass Index (kg/m^2^)

In mutually adjusted models, current income had a significant effect on all outcome scores, while maternal education did not (Supplementary Figure 2B). Finally, linear regression models of SES trajectories showed that outcome scores were influenced by these trajectories, although the effect appears to be driven particularly by current income level (Figure 2B).

## Discussion

Our results are generally consistent with a relationship of lower current and childhood SES with worse functional status and quality of life and more disability. As discussed by Luong *et al*,(1) both socioeconomic behavioral patterns and – more importantly – harmful exposures associated with lower SES through the life course may explain the worse functional status and disability scores, while psychosocial factors may add on to these to influence quality of life.(11) Social-to-biological pathways, such as increased systemic inflammation amongst people of lower SES(20) may well play a role in explaining our results, which are also in line with studies reporting more musculoskeletal pain and generally worse quality of life with decreasing SES.

Important strengths of our study include being, to our knowledge, the first study to examine the relationship between childhood SES and OA outcomes in a Latin American context, and one of few studies to do so worldwide. Major limitations include its cross-sectional nature, small sample size and inherent limitations of measuring a complex construct such as Socioeconomic Status, forcing gross approximations through proxy variables such as income, education and occupation type. Similarly, we were limited by the small number of people in the lowest income groups included in the study, which was due to the fact that our Social Security clinic serves only the families of people with formal employment in a middle to upper-middle class neighborhood.

Taken together with previously mentioned studies showing associations of lower childhood and current SES with higher prevalence and worse outcomes of OA, our study adds to the evidence base for social determinants of health theory and the life course approach within it. While further research is needed to elucidate the influence of socioeconomic life-course trajectories on both OA incidence and outcomes, and the pathways that mediate this relationship, we believe this study joins others in calling for action on social determinants to reduce health disparities.

## Key Messages

### What is already known about this subject?

Socioeconomic Status (SES) is an important contributor to osteoarthritis (OA) prevalence and outcomes. This has been primarily shown in developed countries.

### What does this study add?

Current lower SES, as measured by income, education and occupation type is associated with worse functional status and quality of life and more disability amongst patients with knee and/or hip OA. Childhood SES is associated with worse quality of life amongst these patients, in part mediated through current SES. Ours study adds evidence from a Latin American context.

### How might this impact on clinical practice or future developments

Our results indicate that awareness of patients’ life course socioeconomic status could help reach those at risk for worse outcomes, as well as the need for policy interventions that reduce economic disparities.

## Data Availability

All data produced in the present study are available upon reasonable request to the authors. The R code for the statistical analysis can be found online

https://github.com/julpisanty/OA/blob/main/SES-OA-github.R

## Ethics statement

The study was approved by the local research board under protocol number R-2019-3605-011.

## Data availability statement

Data are available upon reasonable request to the authors. The R code for the statistical analysis can be found at https://github.com/julpisanty/OA/blob/main/SES-OA-github.R

## Patient and public involvement statement

Patients and/or the public were not involved in the design, or conduct, or reporting or dissemination plans of this research.

