## Supplementary Material for "Associations of current and childhood socioeconomic status and health outcomes amongst patients with knee or hip osteoarthritis in a Mexico City family-practice setting"

**Supplementary Figure 1. WOMAC, AMICAL and HAQ-DI Scores by childhood SES varia**

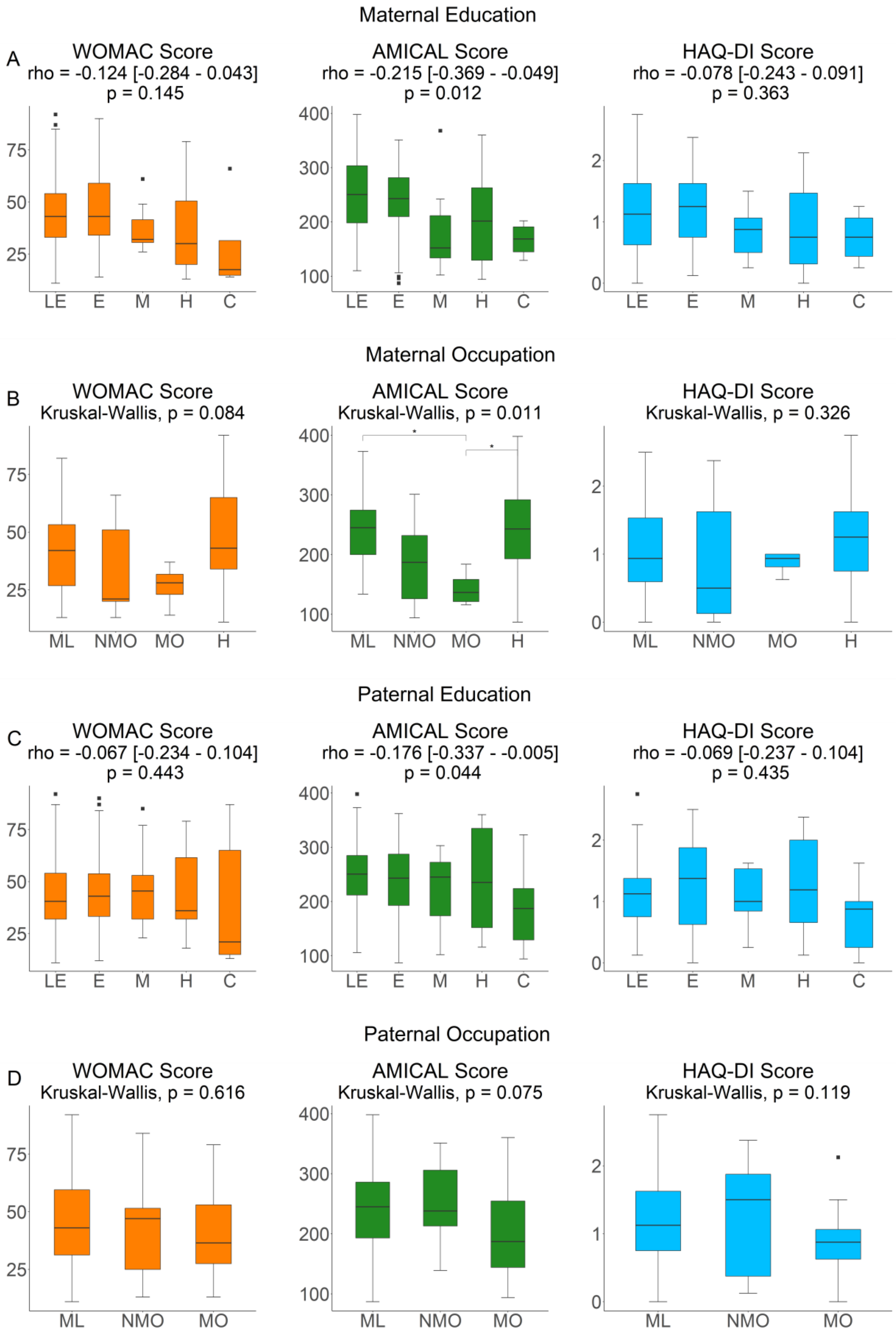


Outcome scores by *A)* Maternal Education *B)* Maternal Occupation type *C)* Paternal Education and *D)* Paternal Occupation type. Spearman analysis for trend was conducted for *A* and *C*. Kruskall-Wallis testing was conducted for *B* and *D*, with post-hoc Dunn tests with Bonferroni correction.

WOMAC: Western Ontario and MacMaster Universities Osteoarthritis Index; AMICAL: Osteoarthritis of Knee and Hip and Quality of Life; HAQ-DI: Stanford Health Assessment Questionnaire Disability Index.

For education level, LE: less than elementary (<6 yr), E: elementary (6 years), M: Middle-school (9 years), H: High-school (12 years), C: College-level or higher (>12 years). For occupation type, H: Home, ML: Manual Labor, NMO: Non-managerial office work, MO: Managerial office work.

*Significant at the p<0.05 level; ** Significant at the p<0.01 level

### Supplementary Figure 2. Linear regression models of effect of maternal education on WOMAC, AMICAL and HAQ-DI scores

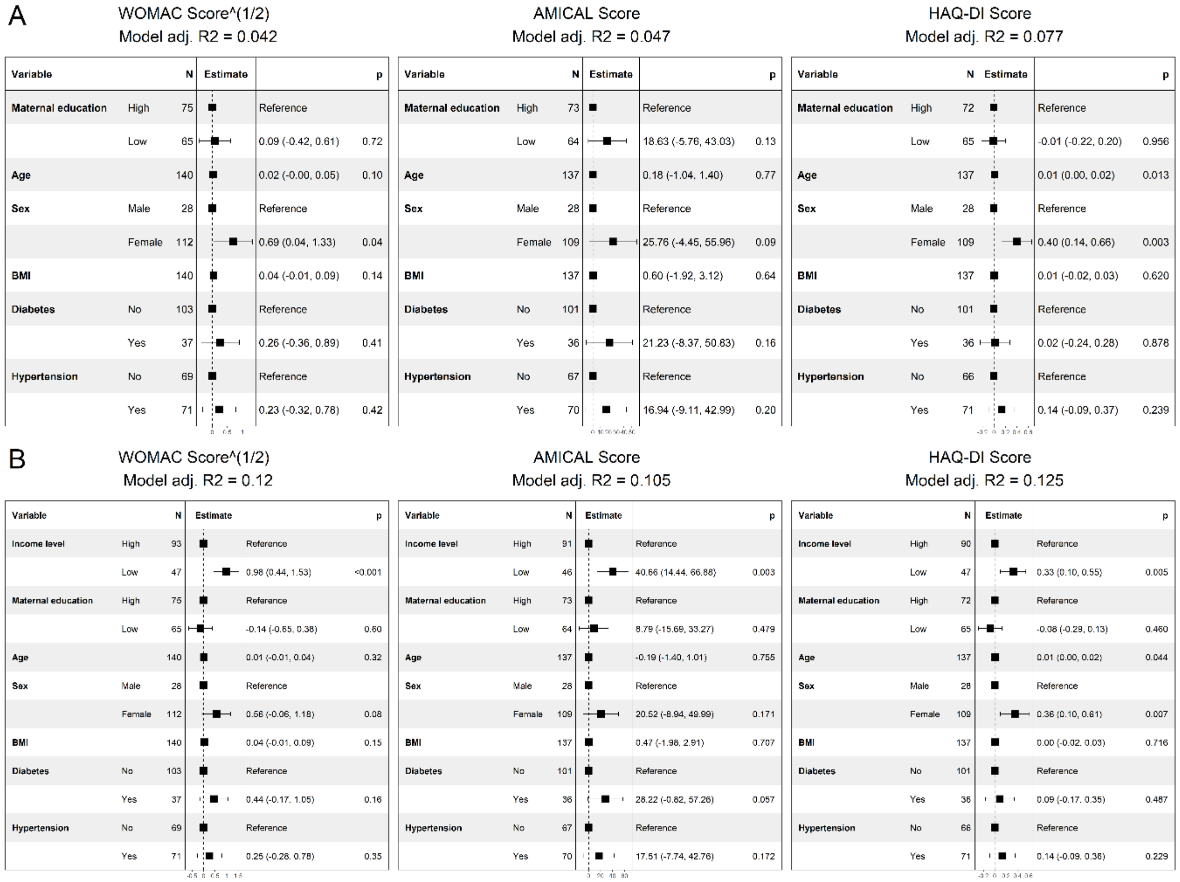


A) Beta coefficients of regression models only including maternal education level.
B) Beta coefficients of regression models including both maternal education level and current income level

WOMAC: Western Ontario and MacMaster Universities Osteoarthritis Index; AMICAL: Osteoarthritis of Knee and Hip and Quality of Life; HAQ-DI: Stanford Health Assessment Questionnaire Disability Index; BMI: Body Mass Index ($kg/m^2$)
